# Metabolic profiling reveals heterogeneity within childhood obesity phenotypes

**DOI:** 10.64898/2026.10.08.26365018

**Authors:** Camilla Guerrini, Mahsa Naghavi Sheikholeslami, Sara Samino, Núria Amigó, Natalia Ferré, Verònica Luque, Maria Vinaixa, Joaquín Escribano, Noelia Ramírez

## Abstract

**Background & Aims:** Childhood obesity is a complex, heterogeneous disease. Children with obesity are often grouped into metabolically healthy obesity (MHO) and metabolically unhealthy obesity (MUO) phenotypes, although this classification may oversimplify the underlying metabolic diversity. We aimed to characterise the metabolic profiles associated with the conventional MHO/MUO classification and to identify data-driven metabolic profiles in 267 children and adolescents with obesity.

**Methods:** In this cross-sectional study, we used serum ^1^H-NMR spectroscopy to quantify lipoproteins, glycoproteins, low-molecular-weight metabolites, and lipid fractions. Univariate comparisons and partial least squares discriminant analysis (PLS-DA) were used to characterise NMR-derived variables associated with MHO and MUO groups. We applied unsupervised k-means clustering based on principal component scores to identify data-driven metabolic profiles independent of the predefined clinical phenotypes.

**Results:** Forty-one NMR-derived variables differed between MUO and MHO (FDR q-value < 0.05), predominantly in the triglyceride-rich lipoprotein and glycoprotein profiles, of which 31 were elevated in MUO. PLS-DA discriminated between MHO and MUO (AUC = 0.84, Q² = 0.33; permutation p < 0.001). Unsupervised clustering identified two data-driven metabotypes that only partially overlapped with the clinical phenotypes: 27% of children with MHO clustered within the adverse metabolic profile (Cluster 2), characterised by a triglyceride-rich and inflammation-associated profile, whereas 41% of children with MUO were assigned to the comparatively favourable profile (Cluster 1). Clustering further revealed additional differences in lipoprotein subclasses and lipid composition that the MHO/MUO classification did not capture.

**Conclusions:** The conventional MHO/MUO classification captures an important component of metabolic variation in childhood obesity but does not fully capture cardiometabolic heterogeneity. Quantitative ¹H-NMR metabolomics identified putative metabotypes characterised by differences in lipoprotein composition and cholesterol distribution beyond clinical phenotypes. These findings support the potential value of metabolomic profiling to refine cardiometabolic risk stratification in children with obesity; however, the relevance of these data-driven metabotypes requires confirmation in independent cohorts and longitudinal follow-up studies.

## Introduction

Obesity is increasingly recognised as a complex, chronic, and heterogeneous disease resulting from the interplay of genetic, environmental, behavioural, socioeconomic, and metabolic factors, rather than a uniform condition attributable solely to unhealthy dietary patterns and insufficient physical activity [1,2]. This multifactorial nature is clinically significant, as individuals with comparable adiposity levels can have markedly different health outcomes [3]. Because conventional BMI-based classifications do not adequately reflect this biological heterogeneity, metabolically healthy obesity (MHO) and metabolically unhealthy obesity (MUO) have been recognised as distinct clinical phenotypes [4]. Since its recognition in the early 1980s, several operational definitions have been proposed (Table S1), with a harmonised definition for children and adolescents established in a 2018 consensus statement [5]. Under this definition, MHO is characterised by the absence of cardiometabolic risk factors, whereas MUO is defined by the presence of at least one abnormal cardiometabolic component, including elevated blood pressure, impaired glucose metabolism, low HDL-cholesterol, or hypertriglyceridaemia [5]. Individuals with MHO meet the BMI criteria for obesity but do not exhibit the cardiometabolic abnormalities typically associated with excess adiposity. Compared with metabolically unhealthy obesity, they generally exhibit lower visceral fat accumulation and a more favourable inflammatory profile [6]. Although clinically useful, this classification relies on predefined thresholds of a limited number of variables. Together with the lack of a consensus definition, this has resulted in considerable variability in the reported prevalence of the two phenotypes, and offers only a partial picture of the complex metabolic profile underlying obesity. Growing evidence suggests that MHO may represent a transitional state preceding the development of cardiometabolic disease within 5–10 years [7,8]. The instability of the MHO phenotype over time and the substantial proportion of individuals reclassified as having MUO within only a few years indicates that this dichotomous classification may not fully capture the aetiology of metabolic risk in childhood obesity [9]. Understanding this heterogeneity is therefore essential to improve risk stratification and inform early interventions to reduce the cardiometabolic, autoimmune, psychosocial, and cognitive consequences of childhood obesity, which may persist into adulthood[10].

Metabolomics has emerged as a powerful approach for characterising disease-associated metabolic alterations, offering the potential to move beyond threshold-based clinical criteria towards more biologically informed metabolic phenotyping. Evidence on the metabolic differences between MHO and MUO has been derived predominantly from adult populations, with comparatively few studies conducted in children and adolescents[11]. Paediatric studies using chromatography-based platforms have reported higher circulating concentrations of branched-chain amino acids, acylcarnitines, and glycerophospholipids, alongside lower concentrations of free fatty acids, in children with MUO than in those with MHO [12,13]. Among available platforms, proton nuclear magnetic resonance (^1^H-NMR) spectroscopy offers high reproducibility, minimal sample preparation, and non-destructive analysis, making it particularly suitable for large-scale clinical and epidemiological studies [14–16]. To the best of our knowledge, only one study has specifically applied ^1^H-NMR to investigate these phenotypes in children, reporting lower lipid and lactate concentrations and higher concentrations of several amino acids and choline-containing phospholipids in metabolically healthy children [17]. Nevertheless, the metabolic signatures underlying cardiometabolic health and dysfunction in childhood obesity remain incompletely characterised.

The present study therefore aims to characterise the serum ¹H-NMR profiles of 267 children and adolescents with obesity, describe metabolic variation associated with the clinically defined MHO and MUO phenotypes, and explore data-driven metabolic profiles beyond this conventional classification. By characterising metabolic variation within and between clinical phenotypes, this study aims to improve our understanding of obesity heterogeneity and to provide a basis for evaluating the prognostic and clinical relevance of metabolic profiles in longitudinal and intervention studies.

## Material & Methods

### Study design

This cross-sectional observational study used data from the baseline visit of the Obemat2.0 clinical trial in primary care (registered at clinicaltrials.gov as NCT03749200) [18,19]. Together with the trial, a registered biobank of biological samples (COLOBEPED, reference C.0004585) was established, providing the infrastructure and biospecimens for the EXPOSOME project (PI22/00852: “EXPOSOME: Identifying the metabolome of childhood obesity and the correlation with the personal exposome”), funded by the Instituto de Salud Carlos III (ISCIII). To limit measurement bias, all participants were assessed with the same anthropometric, blood pressure and blood sampling protocols, and serum samples were processed and stored under identical conditions and analysed on a single NMR platform with automated, standardised quantification. Moreover, NMR acquisition and quantification were performed in a randomised, blinded manner with respect to participants’ MHO/MUO status.

### Study population

The present study included all 267 children and adolescents with obesity (145 males and 122 females) aged 8-14 years, enrolled in Obemat 2.0 between June 2016 and March 2018 during routine health check-ups at the primary healthcare centres in the Camp de Tarragona area (Spain).We defined obesity status as a BMI at or above the 97^th^ percentile, following the criteria established by Hernandez et al., as recommended by the Clinical Practice Guidelines for the Prevention and Treatment of Childhood and Adolescent Obesity of the Spanish National Health System [20,21]. Exclusion criteria included eating disorders, endocrine disorders (such as growth hormone disorders, hypothyroidism, Cushing’s disease or precocious puberty), inability to attend scheduled visits, and concurrent participation in another clinical trial.

### Clinical and anthropometric assessment

Body weight and standing height were measured with a digital scale (SECA 769, precision 50g) and a wall-mounted stadiometer (SECA 216, precision 1 mm), respectively. Waist circumference was measured at the end of expiration, midway between the iliac crest and lower rib using a Holtain non-extensible tape (precision 0.1 cm). BMI (kg/m^2^) was calculated as weight divided by height squared (kg/m^2^), and age- and sex-adjusted BMI z-scores were calculated using WHO references [22,23]. Body fat mass and fat-free mass were assessed by air displacement plethysmography (Bod Pod®) using adjustments for children and adolescents with obesity (COSMED, Life Measurements, Inc, Concord, CA). Systolic (SBP) and diastolic blood pressure (DBP) were measured in mmHg using a Dinamap Pro 100 device on the left arm, with the child seated and the arm resting, at least 20 minutes after arrival. The mean of two measurements taken 5 to 10 min apart was reported. SBP and DBP were standardised as percentiles according to the Clinical Practice Guideline for Screening and Management of High Blood Pressure in Children and Adolescents [24].

### Biochemical analysis and parameters

Blood samples were collected in ethylenediaminetetraacetic acid (EDTA) tubes under overnight fasting conditions. Triglycerides (mg/dL), total cholesterol (mg/dL), low-density lipoprotein cholesterol (LDL-C) (mg/dL), high-density lipoprotein cholesterol (HDL-C) (mg/dL), and glucose (mg/dL) were determined at certified reference-hospital laboratories using routine clinical diagnostic enzymatic methods, while insulin (μIU/mL) was quantified by immunoradiometric assays. The Homeostasis Model Assessment of Insulin Resistance (HOMA-IR) was calculated as [25]:

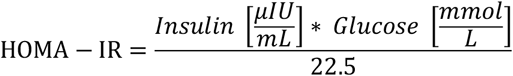

### Clinical cardiometabolic assessment and phenotypes

Children were classified as having metabolically healthy obesity (MHO) or metabolically unhealthy obesity (MUO) adapted from the consensus-based criteria proposed by Damanhoury et al. [5] for children and adolescents. Under these criteria, in line with the American Academy of Paediatrics, children were classified as MUO if they presented at least one of the following five cardiometabolic risk factors: systolic and diastolic blood pressure ≥90^th^ percentile, HOMA-IR ≥90^th^ percentile (z-score 1.2816 from Stavnsbo), HDL cholesterol ≤40mg/dL, or triglycerides ≥110mg/dL [26–28]. Children who did not meet any of these criteria were classified as having MHO. Overall, 126 participants were classified as MHO and 141 as MUO.

In addition, we created a continuous cardiometabolic risk score (MetScore) as the sum of the standardised SBP, DBP, triglycerides, HOMA-IR and HDL cholesterol z-scores, the last one multiplied by -1 (as HDL cholesterol is inversely related to cardiometabolic risk), using the references from Stavnsbo et al. [28].

### Metabolomic profiling by ^1^H-NMR analysis

Blood samples were collected under fasting conditions. Samples were left at room temperature for 30 minutes to allow clotting, then centrifuged at 1500 g for 10 minutes at 4 °C. Serum was carefully separated and aliquoted into 500 μL before being stored at -80 °C until analysis. Samples were processed and stored at the Biobank of the Institut de Recerca Biomèdica Catalunya Sud in Reus and Tarragona (Spain). ^1^H-NMR analyses were performed at Biosfer Teslab. Before analysis, serum samples were thawed overnight at 4 °C and vortexed to homogenise. For each serum sample, 250 μL was diluted with 50 μL of deuterated water and 300 μL of 50 mM phosphate buffer solution (PBS, pH 7.4, containing 30.70 mM Na_2_HPO_4_ and 19.30 mM NaH_2_PO_4_), and then transferred into 5 mm NMR tubes.

High-resolution ^1^H-NMR spectra were acquired on a Bruker Avance III 600 spectrometer (Bruker GmbH, Germany) operating at a proton frequency of 600.20 MHz (14.1 T) and a temperature of 310 K. NMR experiments included 1D Nuclear Overhauser Effect SpectroscopY (NOESY), Carr-Purcell-Meiboom-Gill (CPMG), and diffusion-edited pulse sequences for the characterisation of low- and high-molecular-weight compounds, including lipoproteins and glycoproteins [29,30]. Spectral processing, including phase and baseline correction and peak alignment, was performed automatically using Liposcale^®^ software under standardised conditions to ensure comparability across samples [16].

### Lipoprotein analysis

Lipoprotein analysis was performed using the advanced NMR-based Liposcale assay to quantify lipid concentrations of the main lipoprotein classes (intermediate-density lipoprotein, IDL; very-low-density lipoprotein, VLDL; low-density lipoprotein, LDL; and high-density lipoprotein, HDL), particle concentrations, and particle sizes (Z), as previously described [16,31]. Triglyceride-to-cholesterol ratios (TG/C) were calculated for VLDL, IDL, LDL, and HDL, together with small-to-total particle-number ratios (s/t-P) for VLDL, LDL, and HDL, to further characterise lipoprotein composition and particle distribution.

### Glycoprotein analysis

Circulating glycoprotein-derived N-acetyl signals, resonating between 2.15 and 1.90 ppm, were measured using NOESY experiments, following the procedure described by Fuertes-Martín et al [32,33]. Concentrations of glycoproteins A, B, and F (GlycA, GlycB, and GlycF) were estimated from the total area of three fitted Lorentzian functions [34]. The GlycA area reflects the concentration of protein-bound N-acetylneuraminic acid and N-acetylglucosamine residues; the GlycB area is associated with N-acetylglucosamine residues; and the GlycF area derives from acetyl groups of N-acetylglucosamine, N-acetylgalactosamine, and N-acetylneuraminic acid not bound to proteins. Height-to-width ratios of the GlycA and GlycB signals (H/W-GlycA and H/W-GlycB) were also reported as indices of sugar-protein linkage aggregation state [35].

### Low molecular weight metabolite (LMWM) analysis

Concentrations of nineteen low-molecular-weight metabolites (LMWM) were quantified by deconvoluting and processing CPMG spectra using fully automated software that integrates one-dimensional and two-dimensional J-resolved NMR data, after standard phase and baseline correction and referencing at 2.034 ppm [36]. The nineteen LMWM included: 3-hydroxybutyrate, glycerol, acetate, acetone, alanine, creatinine, creatine, glucose,glutamate, glutamine, glycine, lactate, threonine, proline, histidine, tyrosine, valine, isoleucine, and leucine.

### Lipid analysis

Lipid signal quantification was performed directly from intact serum ^1^H-NMR spectra using an in-house MATLAB-based software developed from the LipSpin platform [37]. Resonance assignments were carried out according to established literature values. Because the area of each NMR resonance is directly proportional to molecular abundance, individual signal areas were converted into molar concentrations by normalisation to external measurements.

The resulting NMR-visible lipid profile comprised a broad range of lipid species, including saturated fatty acids (SFAs), polyunsaturated fatty acids (PUFAs), linoleic acid (LA), docosahexaenoic acid (DHA), arachidonic acid (ARA), eicosapentaenoic acid (EPA), omega-3 (ω-3), omega-6 (ω-6), omega-7 (ω-7), and omega-9 (ω-9) fatty acids, as well as free and esterified cholesterol (FC and EC). In addition, other lipid classes were quantified, including glycerophospholipids (PL), lysophosphatidylcholine (LPC), phosphatidylcholine (PC), and sphingomyelin (SM).

### Ethics

The study was approved by the ethics committees of all participating centres: CEIC Hospital Universitari de Tarragona Joan XXIII (2 March 2016; CP.Obemat 2.0-C.I.01p/2016), CEIC Hospital Universitari Sant Joan de Reus (29 January 2016; 16-01-28/1ass2), and CEIC IDIAP Jordi Gol (26 November 2015; PI14/116), and CEIm Institut d’Investigació Sanitària Pere Virgili (23 February 2023; 032/2023). All participating families provided written informed consent before enrolment in the Obemat2.0 study and for the preservation of blood samples in COLOBEPED, a dedicated registered collection (reference C.0004585). Children aged ≥12 years also provided written assent/consent, as appropriate. The study was conducted in accordance with the Declaration of Helsinki.

### Statistical analysis

Detection frequencies (DFs) were calculated for the variables assessed across the study samples. Parameters with a DF below 70% were excluded from subsequent analyses. Accordingly, 3-hydroxybutyrate (DF = 22%) and glycerol (DF = 36%) were excluded, whereas the remaining NMR-derived variables showed DFs ranging from 76% to 100% and were retained for statistical analysis. Missing values were imputed using half of the minimum non-missing observed value for each variable. Data were then log-transformed to reduce skewness and stabilise variance and z-standardised before multivariate analysis. Clinical variables were available for all participants.

For descriptive reporting, continuous data were summarised on their original scale using the mean and standard deviation (SD), whereas categorical variables were presented as percentages. Differences in continuous clinical and NMR-derived variables between children classified as MHO and MUO were assessed using the Wilcoxon rank-sum test, and categorical variables were compared using the chi-square test. Prior to univariate comparisons, we evaluated age and sex as potential confounders by testing their association with the clinically defined MHO and MUO groups using the Wilcoxon rank-sum test and chi-square test, respectively. Resulting p-values were corrected for multiple testing using the Benjamini–Hochberg false discovery rate (FDR) procedure. NMR-derived variables with FDR-adjusted q-values < 0.05 were considered statistically significant. As neither age nor sex differed significantly between groups (FDR-adjusted q-value = 0.07 and q-value = 0.98, respectively), we used unadjusted analyses for all between-group comparisons.

Partial least squares discriminant analysis (PLS-DA) was applied as a supervised exploratory approach to characterise the multivariable ¹H-NMR profile associated with the clinically defined MHO and MUO groups. The number of latent variables was selected by maximising the cross-validated Q² statistic using 10-fold cross-validation, yielding a final model with two latent variables. Variable Importance in Projection (VIP) scores were calculated to estimate the contribution of each variable to the model, with VIP > 1 considered influential. Group separation was quantified using the area under the receiver operating characteristic curve (AUROC), calculated from out-of-fold predictions, whereas performance relative to random class assignment was assessed using permutation testing (999 permutations) against the null distribution.

We also used unsupervised clustering to explore data-driven metabolic variation in the ¹H-NMR profile, independently of the clinical labels.

Principal component analysis (PCA) was applied to 66 standardised NMR-derived variables, comprising lipoprotein-related measures (n = 30), low-molecular-weight metabolites (n = 17), lipid species (n = 14), and glycoproteins (n = 5), and the first 17 principal components, collectively explaining ≥90% of the total variance, were selected as input to non-hierarchical k-means clustering.

Candidate cluster solutions (k = 2–4) were evaluated using the mean silhouette coefficient, and the final two-cluster solution was selected because it achieved the highest value (mean silhouette=0.30). Because the clusters were generated from the NMR-derived variables, post hoc comparisons of these same variables between clusters were interpreted descriptively as cluster-defining characteristics rather than as independent validation of the clustering solution.

All statistical analyses were performed using MATLAB R2025b (The MathWorks Inc., Natick, MA, USA) with the Statistics and Machine Learning Toolbox [38].

## Results

### Clinical and metabolic parameters of children with MHO and MUO

The anthropometric, clinical, and biochemical characteristics of children classified as having MHO (n = 126) and MUO (n = 141) are presented in Table 1. Age and sex distributions did not differ significantly between the groups. Children with MUO had significantly higher anthropometric and body-composition measures, including higher BMI, body weight, waist-to-height ratio, fat mass index and fat-free mass index, as well as SBP and DBP. We also observed significant differences in their biochemical profiles: participants with MUO had a more adverse lipid profile, with higher total triglyceride concentrationsm and lower HDL-cholesterol concentrations. Greater insulin resistance, as indicated by higher HOMA-IR values, and greater composite cardiometabolic risk, as reflected by a higher MetScore, were consistent with greater metabolic impairment in participants classified as having MUO.

**Table 1.** Anthropometric, clinical, and biochemical characteristics of children with metabolically healthy obesity (MHO) and metabolically unhealthy obesity (MUO). Continuous variables are presented as the mean ± standard deviation (SD), except for systolic and diastolic blood pressure percentiles, which are presented as median (interquartile range, IQR). Sex is reported as number (%). Between-group differences were assessed using the Wilcoxon rank-sum test for continuous variables and the chi-squared test for sex. The resulting p-values were corrected for multiple testing using the Benjamini–Hochberg false discovery rate (FDR) procedure. Bold q-values indicate statistical significance (FDR-adjusted q-values < 0.05).

| Variable | MHO (n=126) | MUO (n=141) | FDR q-values |
| --- | --- | --- | --- |
| **Anthropometric and clinical characteristics** |  |  |  |
| BMI (kg/m<sup>2</sup>) | 24.9 ± 2.2 | 26.5 ± 2.9 | **<0.001** |
| BMI z-score | 2.48 ± 0.4 | 2.66 ± 0.47 | **0.001** |
| Age (years) | 10.6 ± 1.5 | 10.9 ± 1.6 | 0.07 |
| Sex (n, %) |  |  |  |
| Male | 68 (54.0%) | 77 (54.6%) | 0.98 |
| Female | 58 (46.0%) | 64 (45.4%) |  |
| Weight (kg) | 54.5 ± 10.9 | 60.1 ± 13.4 | **0.001** |
| Height (m) | 1.47 ± 0.10 | 1.50 ± 0.10 | **0.04** |
| Waist/Height | 0.56 ± 0.03 | 0.58 ± 0.04 | **<0.001** |
| Fat Mass Index | 9.75 ± 1.85 | 11.0 ± 2.2 | **0.001** |
| Fat Free Mass Index | 15.1 ± 1.8 | 15.5 ± 1.6 | **0.030** |
| SBP (percentile) | 57 (40–75) | 75 (51–94) | **<0.001** |
| DBP (percentile) | 39 (28–55) | 50 (39–70) | **<0.001** |
| **Conventional lipid and glycaemic profile** |  |  |  |
| TC (mg/dL) | 162 ± 28 | 162 ± 29 | 0.95 |
| TG (mg/dL) | 63.7 ± 17.3 | 102 ± 46 | **<0.001** |
| HDL-C (mg/dL) | 57.3 ± 11.1 | 47.6 ± 9.8 | **<0.001** |
| LDL-C (mg/dL) | 92.6 ± 24.8 | 94.2 ± 24.8 | 0.38 |
| Fasting glucose (mg/dL) | 82.8 ± 5.3 | 84.9 ± 5.9 | **0.012** |
| Fasting insulin (μIU /mL) | 9.70 ± 3.34 | 17.8 ± 9.2 | **<0.001** |
| HOMA-IR | 1.99 ± 0.71 | 3.77 ± 2.02 | **<0.001** |
| MetScore | 2.12 ± 1.95 | 6.89 ± 3.08 | **<0.001** |

*Abbreviations:* MHO: metabolically healthy obesity; MUO: metabolically unhealthy obesity; BMI: body mass index; SBP: systolic blood pressure; DBP: diastolic blood pressure; TC: total cholesterol; TG: triglycerides; HDL-C: high-density lipoprotein cholesterol; LDL-C: low-density lipoprotein cholesterol; HOMA-IR: Homeostasis Model Assessment of Insulin Resistance; MetScore: metabolic risk score.

### 1H-NMR serum metabolome profiles distinguishing MHO and MUO

Univariate analysis identified 41 ^1^H-NMR-derived variables that differed significantly between the MUO and MHO groups after FDR correction (q-values < 0.05; **Tables S2**-**S5**). The discriminant variables included lipoprotein-related measures (n = 24), low-molecular-weight metabolites (n = 7), glycoproteins (n = 5), and lipids (n = 5), of which 31 showed higher concentrations in MUO and 10 in MHO. Children with MUO showed higher concentrations of VLDL-related particles (VLDL-P, VLDL-TG, and VLDL-C), triglyceride-enriched HDL particles (HDL-TG), glycoproteins (Glyc-A, Glyc-F), fatty acid measures (ω-9 and ω-6 + ω-7), alanine, lactate, and glutamate. Conversely, MHO was associated with higher HDL-C and larger LDL particle size (LDL-Z), with the latter showing the strongest association with a healthy clinically defined metabolic phenotype. **Figure 1A** shows the concentrations of nine significantly altered compounds.

**Figure 1.**
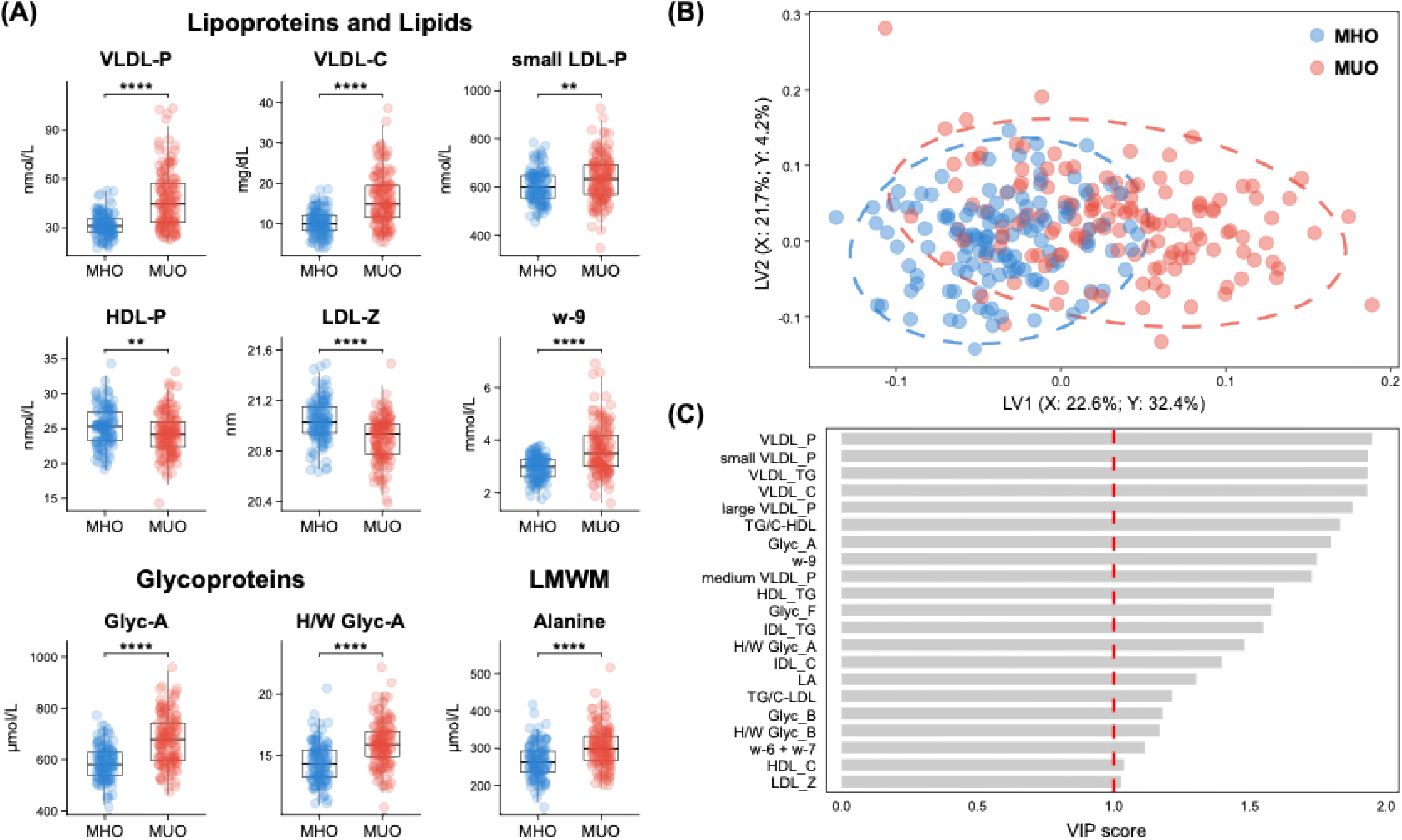
Results of the NMR analysis comparing metabolically unhealthy obesity (MUO) and metabolically healthy obesity (MHO). (A) Box plots show the distribution of nine significantly altered NMR-derived metabolic compounds between MHO (n=126) and MUO (n=141). We assessed statistical significance using the Wilcoxon rank-sum test (** = q-value < 0.01; **** = q-value < 0.0001). (B) PLS-DA plot show the distribution of children according to the clinical metabolic health status (MHO/MUO). (C) Variable Importance in Projection (VIP) scores for the variables contributing to the PLS-DA model, highlighting variables with VIP score > 1.

To characterise the multivariable NMR profile associated with the clinically defined MHO and MUO groups, NMR data were analysed by supervised partial least squares discriminant analysis (PLS-DA) (**Figure 1B**). The model, comprising two latent variables, explained a cumulative 44.3% of the variance in the NMR variables predictor matrix [R²X(cum): LV1 = 22.6%, LV2 = 21.7%] and 36.6% of the variance in class membership [R²Y (cum: LV1=32.4%, LV2=4.2%]. Cross-validated model performance, assessed by 10-fold cross-validation, yielded a Q² of 0.330, and permutation testing (999 permutations) confirmed that the observed group separation exceeded that expected under random class assignment (p-value < 0.001). Among variables with VIP > 1, the 20 variables with the highest VIP scores were predominantly related to triglyceride-rich lipoprotein metabolism and inflammatory processes, including VLDL particle concentration (VLDL-P), VLDL cholesterol and triglycerides (VLDL-C, VLDL-TG), small, medium, and large VLDL subclasses, and the glycoprotein markers GlycA, GlycB, and GlycF (including the GlycA and GlycB height-to-width [H/W] ratios), together with ω-9 fatty acids. Additional contributors included HDL-TG and the TG/C-HDL, IDL-C and IDL-TG, LDL-TG and TG/C-LDL, ω-6+ω-7 fatty acids, and linoleic acid (LA) (**Figure 1C**). ROC analysis based on cross-validated out-of-fold predictions showed cross-validated separation between the clinically defined MUO and MHO groups (AUC = 0.84; **Figure S1**). Because triglycerides and HDL-C contributed to the clinical phenotype definition and were represented directly or indirectly in the NMR profile, this AUC reflects separation of the predefined clinical groups rather than independent diagnostic performance.

### NMR metabolomics-based clustering: metabolic heterogeneity beyond clinical cardiometabolic classification

The MHO/MUO classification relies on discrete, externally defined criteria that may not fully capture the continuous, multidimensional nature of metabolic variation reflected in the ¹H-NMR profile. To explore group structures emerging from intrinsic patterns in the metabolic data rather than predefined labels, we applied unsupervised clustering. This approach identified two data-driven metabotypes: a comparatively favourable metabolic profile (Cluster 1) and a comparatively adverse profile characterised by higher levels of triglyceride-rich lipoprotein and inflammation-associated glycoproteins (Cluster 2). Age, sex, BMI, waist-to-height ratio, fat mass index, fat-free mass index, and blood pressure did not differ significantly between clusters after FDR correction. Cluster 2 had significantly higher clinical concentrations of triglycerides, total cholesterol, VLDL-C, LDL-C, fasting insulin, and HOMA-IR. Post hoc descriptive comparisons identified 51 NMR-derived variables that differed between clusters (FDR-adjusted q-values < 0.05; **Table S6**), compared with 41 variables distinguishing MHO from MUO. Notably, 17 of these cluster-defining variables had not differed between MHO and MUO. Because these variables were used to generate the clusters, their between-cluster differences describe the profiles defining the clustering solution and should not be interpreted as independent associations or validation. Most variables meeting the FDR threshold were elevated in Cluster 2, with a smaller set elevated in Cluster 1. Among the lipid-related variables, EC, FC, PC, SM, LPC, PL, PUFA, ω-3 were higher in Cluster 2. Glycoprotein variables (GlycA, GlycB, and GlycF) were likewise higher in Cluster 2, as were variables related to the VLDL, IDL, and LDL fractions. By contrast, HDL-C did not differ significantly between clusters.

Among low-molecular-weight metabolites, creatinine, tyrosine, valine, and isoleucine were higher in Cluster 2, whereas glutamine was higher in Cluster 1. NMR-derived variables that also differed in the MHO/MUO comparison generally showed larger absolute effect sizes between clusters, as expected because the clustering algorithm was designed to minimise within-cluster variation and maximise separation between the resulting profiles. Lipoprotein particle concentrations followed a similar pattern. VLDL-P and its large, medium, and small subclasses were higher in Cluster 2 than in Cluster 1 (all q-values < 0.001), as were LDL-P and its large, medium, and small subclasses (all q-values < 0.001). For HDL, total HDL-P and small HDL-P were higher in Cluster 2, whereas large and medium HDL-P did not differ significantly between clusters. Average HDL particle size (HDL-Z) was larger in Cluster 1. The concentrations of nine selected NMR-derived variables are presented in **Figure 2A** by metabolic category, lipid-related variables, glycoproteins, lipoproteins, and low-molecular-weight metabolites, illustrating the comparatively adverse lipid and inflammation-associated profile of Cluster 2. Cluster 2 also showed a higher composite cardiometabolic risk score (MetScore) than Cluster 1 (**Figure 2B**; q-value < 0.001), indicating correspondence between the NMR-derived cluster assignment and the overall clinical cardiometabolic profile.

**Figure 2.**
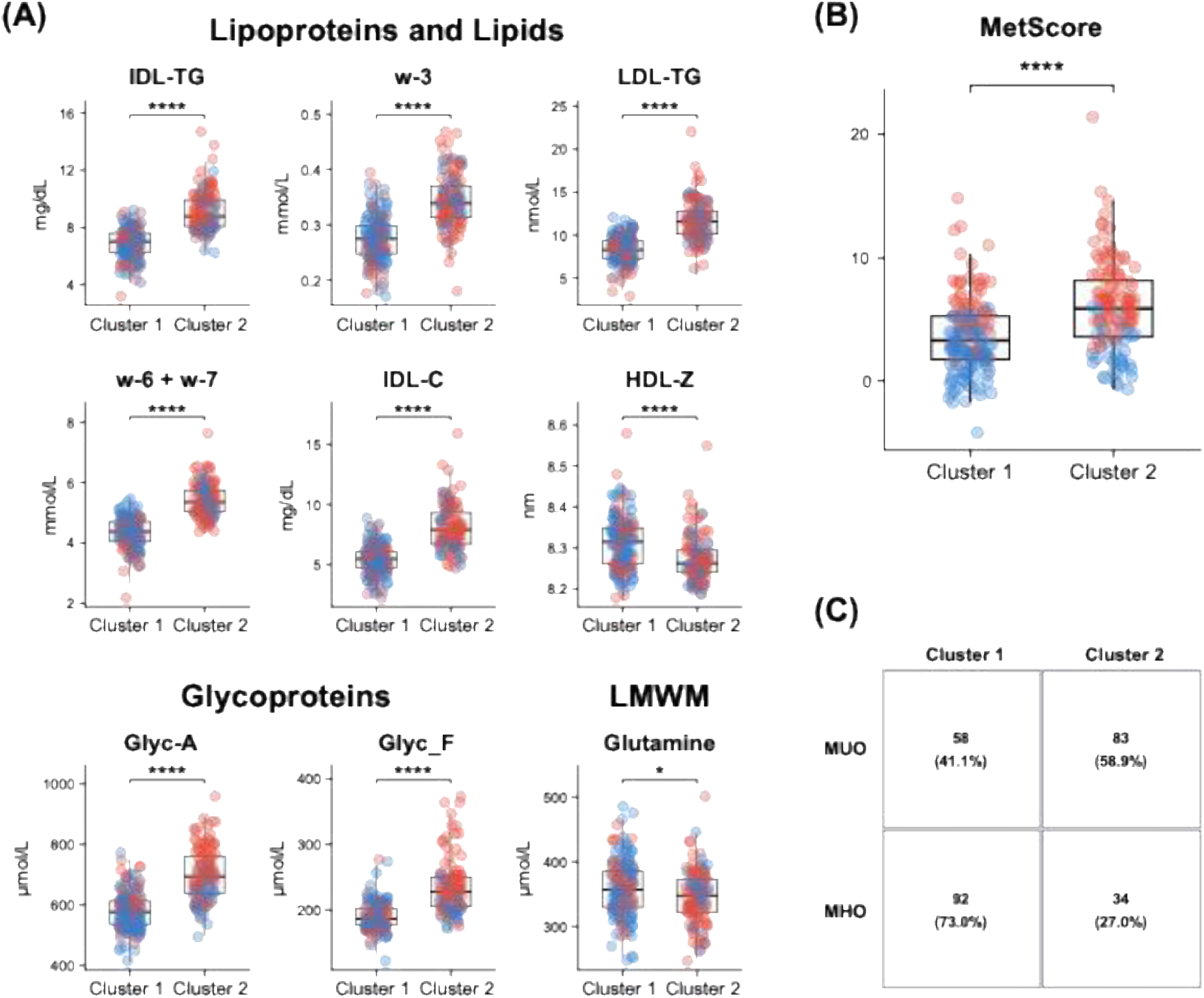
Results of the unsupervised k-means clustering analysis. (A) Box plots showing the distribution of nine significantly altered NMR-derived metabolic compounds between Cluster 1 (n=150) and Cluster 2 (n=117). (B) Cardiometabolic risk score (MetScore) by NMR-derived metabolic cluster. Box plot shows the distribution of the composite MetScore in clusters derived from unsupervised k-means clustering of serum NMR metabolomic profiles. We assessed statistical significance using the Wilcoxon rank-sum test (** = q-value < 0.01; **** = q-value < 0.0001). (C) Confusion matrix comparing the clinical MHO/MUO classification with NMR-derived k-means clusters. Rows represent the clinical classification (MUO, MHO) and columns represent the metabolic cluster assignment (Cluster 1, Cluster 2) derived from NMR metabolomic profile. Values indicate the number and percentage of children in each category.

To assess the correspondence between the NMR-derived clusters and the conventional MHO/MUO classification, we constructed a confusion matrix (**Figure 2C**). Of the 126 children with MHO, 92 (73%) were assigned to Cluster 1 and 34 (27%) to Cluster 2, whereas, of the 141 children with MUO, 58 (41%) were assigned to Cluster 1 and 83 (59%) to Cluster 2. The two classifications therefore showed partial correspondence, with children with MHO predominating in Cluster 1 (61%) and those with MUO predominating in Cluster 2 (71%). However, discordant assignments were frequent: 27% of children with MHO were assigned to the comparatively adverse metabolic profile (Cluster 2), whereas 41.1% of those with MUO were assigned to the comparatively favourable profile (Cluster 1). These findings demonstrate incomplete correspondence between the clinical classification and the data-driven NMR profiles, highlighting metabolic heterogeneity within both clinical phenotypes.

## Discussion

In this cohort of 267 children and adolescents with obesity, quantitative metabolic profiles showed differences between the conventional MHO/MUO phenotypes, particularly in triglyceride-rich lipoprotein and inflammation-associated glycoprotein signatures, capturing only part of the metabolic heterogeneity within this population. Unsupervised clustering identified two data-driven metabotypes that were discordant with the clinical phenotype in 27% of children with MHO and 41% of children with MUO, indicating that the clinical label and the underlying metabolic profile frequently diverge.

### Metabolic patterns of the clinical MHO/MUO phenotypes

Consistent with its clinical definition, MUO had elevated VLDL particles and VLDL triglycerides, smaller LDL particle size, lower HDL cholesterol, and higher glycoprotein variables associated with low-grade inflammation (GlycA, GlycB, and GlycF). Overall, these findings suggest that MUO is characterised by a coordinated pattern of dyslipidaemia and low-grade inflammation.

However, because triglycerides and HDL-C contributed to the MHO/MUO definition and were represented directly or indirectly in the NMR metabolic profile, the PLS-DA performance should not be interpreted as evidence of independent diagnostic value. Rather, the supervised analysis provides a higher-resolution characterisation of the lipoprotein particle distribution and glycoprotein alterations accompanying the conventional clinical classification. These findings agree with previous evidence linking insulin resistance, dyslipidaemia, and chronic low-grade inflammation to adverse cardiometabolic outcomes in children with obesity [10,39]. However, the clinical classification only partially captured the variation observed in the metabolic profiles.

### Data-driven metabotypes beyond the clinical MHO/MUO classification

When metabolic profiles were analysed independently of the clinical MHO/MUO classification, two well-distinguished data-driven metabotypes emerged. One was characterised by a more favourable cardiometabolic pattern, with lower concentrations of triglyceride-rich lipoproteins and inflammation-associated glycoproteins (Cluster 1), whereas the other showed a more adverse profile, marked by elevated triglyceride-rich lipoproteins, low-grade inflammation, and differences in LDL and HDL particle composition (Cluster 2). Although MUO was characterised by a triglyceride-rich lipoprotein and low-grade inflammation-associated profile, unsupervised clustering identified additional variation in lipoprotein composition and cholesterol distribution. Interestingly, a substantial proportion of children were assigned to a metabotype inconsistent with their clinical phenotype. Some children classified as MHO exhibited a metabolomic profile resembling that of MUO, whereas others classified as MUO showed a comparatively favourable metabolic profile. The identification of an adverse metabolic profile in a subset of children classified as MHO raises the possibility that these children may be at greater risk of future metabolic deterioration and may represent a transient state preceding the development of MUO phenotype. These findings suggest that childhood obesity comprises multiple interconnected metabolic dimensions that are only partially reflected by conventional clinical and biochemical criteria used in current MHO/MUO definitions. However, this hypothesis **warrants longitudinal validation.**

### Comparison with previous studies

Our findings are consistent with emerging evidence indicating that metabolic heterogeneity in childhood obesity extends beyond traditional risk-factor definitions. In the Generation R Study, Schipper et al. demonstrated that selected metabolites improved the identification of children with an unhealthy metabolic phenotype, using targeted mass-spectrometry-based metabolomics [40]. Together with our results, these observations suggest that metabolomic profiling may provide additional resolution of biological variation within conventional cardiometabolic classifications.

Few studies have specifically applied ¹H-NMR spectroscopy to investigate differences between paediatric MHO and MUO phenotypes. A previous study reported lower lipid and lactate concentrations and higher concentrations of several amino acids and phosphocholines in metabolically healthy children, consistent with the comparatively adverse lipid and lactate profile observed here in MUO [17]. The present study extends these observations by including a larger cohort and using a quantitative NMR platform capable of resolving lipoprotein particle concentrations, sizes, and subclasses. In addition, the combination of supervised and unsupervised multivariate methods allowed us to assess metabolic differences between the clinically defined MHO and MUO groups and to examine its correspondence with exploratory data-driven metabolic profiles.

Complementary evidence from mass spectrometry-based metabolomics in children and adolescents with obesity identified differences in fatty acids, amino acids, and glucose-related metabolites between MHO and MUO. The greater prominence of lipoprotein and glycoprotein variables in the present study probably reflects differences in analytical coverage rather than a contradictory biological pattern. NMR is particularly well suited to quantitative profiling of lipoprotein particle concentrations, sizes, and subclasses, as well as glycoproteins. However, NMR’s high detection limit restricts the assessment of low-abundance metabolites to a smaller range of compounds, such as branched-chain and aromatic amino acids, glucose, lactate, and ketone bodies. On the other hand, mass-spectrometry-based platforms, typically coupled to liquid or gas chromatography, reaches nanomolar or lower sensitivity, providing greater sensitivity and broader coverage of low-abundance metabolites. These include amino acids and their derivatives, acylcarnitines across a broad range of chain lengths, and specific lipid species such as phosphatidylcholines, sphingomyelins, ceramides, and triacylglycerols[41]. These analytical approaches are therefore complementary, and their integration could provide a more comprehensive characterisation of metabolic health in childhood obesity [12,15,42].

### Biological interpretation of the metabolomic profiles

Glycoprotein variables may be particularly relevant to the inflammatory component of metabolic health. GlycA, reflecting circulating acute-phase glycoproteins, has been associated with adverse cardiovascular risk profiles and vascular inflammation [33,43]. Longitudinal data from the Avon Longitudinal Study of Parents and Children and the Cardiovascular Risk in Young Finns Study have shown that GlycA, but not C-reactive protein (CRP), predicted incident hypertension and metabolic syndrome over 9–10 years of follow-up [33]. These findings support further investigation of GlycA and related glycoprotein signals as prospective markers of cardiometabolic risk. However, the present cross-sectional study cannot establish whether GlycA, GlycB, or GlycF has anticipatory or incremental predictive value in children with obesity. Their addition to paediatric risk-prediction models should therefore be tested against conventional clinical variables in independent longitudinal cohorts.

A plausible biological framework for the co-occurrence of triglyceride-rich lipoprotein abnormalities and glycoprotein-associated inflammation involves insulin resistance and visceral adiposity. In insulin-resistant states, increased adipose-tissue lipolysis and fatty-acid flux to the liver, together with increased hepatic de novo lipogenesis, can promote hepatic VLDL production and secretion, while impaired lipoprotein-lipase-mediated clearance favours the accumulation of triglyceride-rich lipoproteins [44,45]. CETP-mediated exchange of triglycerides and cholesteryl esters between VLDL, LDL, and HDL generates triglyceride-enriched LDL and HDL particles, which are subsequently remodelled by hepatic lipase into smaller, denser LDL particles and smaller, cholesterol-depleted HDL particles—the characteristic atherogenic lipoprotein phenotype [45]. This mechanism is compatible with the higher VLDL triglyceride and particle concentrations, smaller LDL particle size, and lower HDL cholesterol observed in MUO. The cluster-derived profiles showed a related but not identical pattern: Cluster 2 had higher VLDL and LDL particle concentrations and a smaller average HDL particle size, whereas LDL particle size and HDL-C did not differ significantly between clusters.

In parallel, insulin resistance and central adiposity are associated with low-grade systemic inflammation and altered hepatic production and glycosylation of acute-phase proteins, which may contribute to the higher GlycA, GlycB, and GlycF signals observed in MUO and Cluster 2. The co-occurrence of these lipoprotein and glycoprotein patterns is therefore biologically plausible, but the present data cannot determine whether they share a causal pathway or whether insulin resistance or visceral adiposity mediates their association. Strengths and limitationsThe strengths of this study include its comparatively large paediatric cohort, quantitative characterisation of lipoprotein particles and glycoprotein signals, and complementary use of clinically defined and data-driven analytical approaches. Nevertheless, these results should be interpreted in light of several limitations. First, the cross-sectional design does not allow us to determine whether the NMR-derived clusters predict future cardiometabolic outcomes or represent stable metabolic profiles over time. Second, the cohort was recruited from a single Mediterranean region, and we did not test the clustering solution in an external validation cohort, limiting its generalisability. Third, we did not account for pubertal stage, which strongly influences lipoprotein metabolism and insulin sensitivity at these ages, and we did not adjust between-group comparisons for potential confounders. Fourth, we recruited participants in primary care, which may not represent all children with obesity, and we imputed NMR-derived values below the detection limit as half the minimum observed value, which may bias estimates for less frequently detected variables. Finally, although ¹H-NMR spectroscopy is highly reproducible, it captures a narrower range of low-abundance metabolites than mass-spectrometry-based platforms. This difference in analytical coverage may partly explain why our discriminating signature was dominated by lipoprotein and glycoprotein variables rather than by the amino acids and acylcarnitines more commonly reported in MS-based paediatric studies.

### Clinical and research implications

From a clinical and public health perspective, these findings show that the MHO/MUO label does not fully describe the variation present in quantitative NMR profiles within either clinical phenotype. This observation is relevant given evidence that paediatric MHO status is not stable over time [46]. Comparable changes in metabolic status have been reported in adult cohorts, where a substantial proportion of individuals with MHO transition to an unhealthy metabolic phenotype, with a correspondingly increased risk of subclinical atherosclerosis, over several years of follow-up [8]. Longitudinal studies integrating metabolic profiles with clinical risk factors and subsequent cardiometabolic outcomes will be important to determine the clinical relevance of these data-driven profiles and their potential role in personalised monitoring of children with obesity.

## Conclusion and future work

Our findings indicate that cardiometabolic heterogeneity in childhood obesity extends beyond the conventional MHO/MUO classification. Unsupervised clustering of the NMR metabolic profile identified additional metabolic variations in lipoprotein cholesterol distribution and lipid composition that were not fully captured by the clinical phenotypes. From a nutritional and clinical research perspective, these findings support further evaluation of quantitative lipoprotein and glycoprotein profiling as an adjunct to conventional paediatric obesity assessment. Such profiling may provide a more comprehensive characterisation of metabolic health; however, its clinical utility for risk stratification and decision-making requires longitudinal validation.

Future research should focus on validating these findings in independent paediatric cohorts from diverse geographical and ethnic backgrounds and determine whether the identified data-driven metabotypes predict subsequent cardiometabolic deterioration, hepatic steatosis, or response to treatment. More studies should also assess whether specific metabolic features can improve the identification of children at increased risk beyond conventional clinical variables. Finally, intervention studies are needed to establish whether children with different metabolic profiles respond differently to lifestyle or nutritional interventions and, subsequently, whether interventions stratified by these profiles provide greater cardiometabolic benefits than approaches based on BMI or conventional clinical phenotypes.

## Acknowledgments

We acknowledge all the participating families in the Obemat 2.0 study for their dedication and effort, as well as the healthcare centre professionals at primary care centres, and laboratory nurses at Hospital Joan XXIII and Hospital Universitari Sant Joan de Reus. We are also grateful for the support provided by the Biobank IRBCatsud staff. We gratefully acknowledge all the Obemat2.0 Study group:

Closa-Monasterolo R, Escribano J, Luque V, Feliu-Rovira A, Ferré N, Muñoz-Hernando J, Gutiérrez-Marín D, Zaragoza-Jordana M, Gispert-Llauradó M, Rubio-Torrents MC, Núñez-Roig M, Alcázar M, Sentís S, Esteve M (Pediatric Nutrition and Human Development Research Unit, Universitat Rovira i Virgili, IRBCatsud, Reus). Monné-Gelonch R, Basora JM, Flores G, Hsu P, Rey-Reñones C, Alegret C (IDIAP Jordi Gol i Gurina, USR Reus); Guillen N, Alegret-Basora C, Ferre R (Hospital Universitari Sant Joan de Reus); Arasa F (Hospital Verge de la Cinta de Tortosa, Institut Català de la Salut); Alejos AM, Diéguez M, Serrano MA, Mallafré M, González-Hidalgo R, Braviz L, Resa A, Palacios M, Sabaté A, Simón L (Hospital Lleuger de Cambrils, Sagessa); Losilla AC, De La Torre S, Rosell L, Adell N, Pérez C, Tudela-Valls C, Caro-Garduño R, Salvadó O, Pedraza A, Conchillo J, Morillo S (CAP Llibretat, Reus, Institut Català de la Salut); Garcia S, Mur EM, Paixà S, Tolós S, Martín R, Aguado FJ, Cabedo JL, Quezada LG (CAP Marià Fortuny, Reus, Sagessa); Domingo M, Ortega M, Garcia RM, Romero O, Pérez M, Fernández M, Villalobos ME (CAP Salou, Institut Català de la Salut); Ricomà G, Capell E, Bosch M, Donado A, Sanchis FJ, Boix A, Goñi X, Castilla E, Pinedo MM, Supersaxco L, Ferré M, Contreras J (CAP Rambla Nova, Institut Català de la Salut, Tarragona); Sanz-Manrique N, Lara A, Rodríguez M, Pineda T, Segura S, Vidal S, Salvat M (CAP Les Borges del Camp-Montroig del Camp, Institut Català de la Salut); Mimbrero G, Albareda A, Guardia J, Gil S, Lopez M (CAP Sant Pere, Reus, Institut Català de la Salut); Ruiz-Escusol S, Gallardo S (CAP Bonavista-La Canonja, Tarragona, Institut Català de la Salut); Machado P, Bocanegra R (CAP Torreforta-La Granja, Tarragona, Institut Català de la Salut); Espejo T, Vendrell M (ABS Vandellòs-L’Hospitalet de l’Infant, Sagessa); Solé C, Urbano R, Vázquez MT, Fernández-Antuña L (CAP Muralles, Tarragona); Barrio M, Baudoin A, González N (CAP El Morell, Institut Català de la Salut); Olivé R, Lara RM, Dinu C, Vidal C (CAP Sant Pere i Sant Pau, Tarragona, Institut Català de la Salut); González S, Ruiz-Morcillo E, Ainsa ME, Vilalta P, Aranda B (CAP Sant Salvador, Tarragona, Institut Català de la Salut); Boada A, Balcells E (ABS Alt Camp Est, Vilarodona, Institut Català de la Salut).

In collaboration with Biosfer Teslab (Reus, Spain; webpage: https://biosferteslab.com/)

## Author Contribution

Camilla Guerrini: Conceptualisation, Study Design, Data curation, Formal Analysis, Methodology, Visualisation, Manuscript writing - original draft. Mahsa Naghavi Sheikholeslami: Conceptualisation, Data curation, Formal analysis, Visualisation, Writing - original draft. Sara Samino: Data curation, Formal Analysis. Núria Amigó: Data curation, Formal Analysis, Manuscript writing - review & editing. Natalia Ferré: Conceptualisation, Supervision, Resources, Manuscript writing - review & editing. Verònica Luque: Conceptualisation, Supervision, Resources, Manuscript writing - review & editing. Maria Vinaixa: Conceptualisation, Supervision, Writing - review & editing. Joaquín Escribano: Conceptualisation, Funding acquisition, Manuscript writing - review & editing. Noelia Ramírez: Conceptualisation, Funding acquisition, Project administration, Supervision, Validation, Manuscript writing - review & editing.

## Data availability

Data will be available upon request.

