## Supplementary Information for "Metabolic profiling reveals heterogeneity within childhood obesity phenotypes"

### CONTENT

### TABLES

**Table S1. Definitions and classification criteria for metabolically healthy obesity (MHO) and metabolically unhealthy obesity (MUO) in children and adolescents.**

**Table S2. Comparison of serum lipoprotein profiles in children with metabolically healthy obesity (MHO) and metabolically unhealthy obesity (MUO).**

**Table S3. Comparison of glycoprotein variables in children with metabolically healthy obesity (MHO) and metabolically unhealthy obesity (MUO).**

**Table S4. Comparison of low-molecular-weight metabolite (LMWM) profiles in children with metabolically healthy obesity (MHO) and metabolically unhealthy obesity (MUO).**

**Table S5. Comparison of advanced lipid profiles in children with metabolically healthy obesity (MHO) and metabolically unhealthy obesity (MUO).**

**Table S6. Comparison of anthropometric, clinical, biochemical, and NMR-metabolomics variables between the two clusters derived from unsupervised k-means clustering.**

### FIGURES

**Figure S1. Receiver operating characteristic (ROC) curve for the PLS-DA model discriminating MUO from MHO.**

**Table S1. Definitions and classification criteria for metabolically healthy obesity (MHO) and metabolically unhealthy obesity (MUO) in children and adolescents.**

| Study | Age (range) | Criteria for MHO phenotype | Criteria for MUO phenotype | Classification |
|---|---|---|---|---|
| Damanhoury et al. (1) | 2-18 | ▪ SBP or DBP < 90th percentile;<br>▪ TG ≤ 150 mg/dL (or ≤ 1.7 mmol/L);<br>▪ HDL-C > 40 mg/dL (or > 1.03 mmol/L);<br>▪ a measure of glycemia | ▪ SBP or DBP ≥ 90th percentile;<br>▪ TG ≥ 150 mg/dL (or ≥ 1.7 mmol/L);<br>▪ HDL-C ≤ 40 mg/dL (or ≤ 1.03 mmol/L);<br>▪ a measure of glycemia. | MHO, 0 risk factors<br>MUO, ≥ 1 risk factor |
| Prince et al. (2) | 8–17 | Method a: HOMA-IR score < 3.16<br>Method b:<br>▪ SBP or DBP < 90th percentile for age, sex, and height;<br>▪ TG < 1.25 mmol/L;<br>▪ HDL-C > 1.02 mmol/L;<br>▪ fasting glucose < 5.6 mmol/L | Method a, HOMA-IR score ≥ 3.16<br>Method b:<br>▪ SBP or DBP ≥ 90th percentile for age, sex, and height;<br>▪ TG ≥ 1.25 mmol/L;<br>▪ HDL-C ≤ 1.02 mmol/L;<br>▪ fasting glucose ≥ 5.6 mmol/L | MHO, 0 risk factors<br>MUO, ≥ 1 risk factor |
| Camhi et al. (3) | 8–18 | ▪ SBP or DBP < 90th percentile for age, sex, and height;<br>▪ TG < 110 mg/dL;<br>▪ HDL-C > 40 mg/dL; glucose < 100 mg/dL | ▪ SBP or DBP ≥ 90th percentile for age, sex, and height; TG ≥ 110 mg/dL; HDL-C ≤ 40 mg/dL; fasting glucose ≥ 100 mg/dL | MHO, < 2 risk factors<br>MUO, ≥ 2 risk factors |
| Mangge et al. (4) | 8–18 | ▪ waist circumference < 90th percentile;<br>▪ TG < 1.7 mmol/L;<br>▪ HDL-C ≥ 1.03 mmol/L in males or ≥ 1.29 mmol/L in females;<br>▪ fasting glucose < 5.6 mmol/L | ▪ waist circumference ≥ 90th percentile;<br>▪ TG ≥ 1.7 mmol/L;<br>▪ HDL-C < 1.03 mmol/L in males or < 1.29 mmol/L in females;<br>▪ fasting glucose ≥ 5.6 mmol/L | MHO, < 3 risk factors<br>MUO, ≥ 3 risk factor |
| Vukovic et al. 36(5) | 6–19 | ▪ Insulin-sensitive (ISG) ≤ 2.75 | ▪ Insulin-resistant (IRG) ≥ 6.16 | - |

*Abbreviations:* SBP, systolic blood pressure; DBP, diastolic blood pressure; TG, triglycerides; HDL, high-density lipoprotein cholesterol; HOMA-IR, Homeostasis Model Assessment of Insulin Resistance.

**Table S2. Comparison of serum lipoprotein profiles of children with metabolically healthy obesity (MHO) and metabolically unhealthy obesity (MUO).** Data are presented as the mean ± standard deviation (SD). Wilcoxon rank-sum test was used to compare groups. The resulting *p*-values were corrected for multiple testing using the Benjamini–Hochberg false discovery rate (FDR) procedure. Bold values indicate statistical significance (FDR-adjusted q-value < 0.05).

| Lipid and lipoprotein variables | MHO (n=126) | MUO (n=141) | FDR q-values |
|---|---|---|---|
| ***Total lipids*** | | | |
| Total cholesterol (mg/dL) | 176 ± 19 | 176 ± 22 | 0.91 |
| Total triglycerides (mg/dL) | **71.6 ± 12.5** | **96.9 ± 30.1** | **<0.001** |
| ***Lipoprotein lipid distribution*** | | | |
| VLDL-C (mg/dL) | 10 ± 3 | 16± 6 | **<0.001** |
| VLDL-TG (mg/dL) | 43.6 ± 9.9 | 65.2 ± 24.9 | **<0.001** |
| TG/C-VLDL | 4.46 ± 0.77 | 4.18 ± 0.56 | **0.005** |
| IDL-C (mg/dL) | 5.98 ± 1.66 | 7.14 ± 2.19 | **<0.001** |
| IDL-TG (mg/dL) | 7.31 ± 1.30 | 8.36 ± 1.80 | **<0.001** |
| TG/C-IDL ratio | 1.26 ± 0.18 | 1.21 ± 0.17 | **0.01** |
| LDL-C (mg/dL) | 108 ± 16 | 105 ± 17 | 0.39 |
| LDL-TG (mg/dL) | 9.39 ± 2.04 | 10.1 ± 2.1 | **0.03** |
| TG/C-LDL ratio | 0.09 ± 0.01 | 0.10 ± 0.03 | **<0.001** |
| HDL-C (mg/dL) | 51.9 ± 6.1 | 47.1 ± 5.4 | **<0.001** |
| HDL-TG (mg/dL) | 11.2 ± 1.81 | 13.2 ± 3.08 | **<0.001** |
| TG/C-HDL ratio | 0.22 ± 0.04 | 0.28 ± 0.07 | **<0.001** |
| ***Lipoprotein size distribution*** | | | |
| VLDL-Z diameter (nm) | 42.2 ± 0.19 | 42.2 ± 0.20 | **0.03** |
| VLDL-P (nmol/L) | 31.6 ± 7.06 | 47.5 ± 17.8 | **<0.001** |
| LargeVLDL-P (nmol/L) | 0.99 ± 0.24 | 1.38 ± 0.42 | **<0.001** |
| MediumVLDL-P (nmol/L) | 3.32 ± 1 | 4.85 ± 2.29 | **<0.001** |
| SmallVLDL-P (nmol/L) | 27.3 ± 6.11 | 41.3 ± 15.5 | **<0.001** |
| LDL-Z diameter (nm) | 21 ± 0.17 | 20.9 ± 0.17 | **<0.001** |
| LDL-P (nmol/L) | 1053 ± 147 | 1056 ± 172 | 0.62 |
| LargeLDL-P (nmol/L) | 180 ± 23 | 169 ± 22.1 | **<0.001** |
| MediumLDL-P (nmol/L) | 268 ± 73.9 | 254 ± 75.2 | 0.32 |
| SmallLDL-P (nmol/L) | 604 ± 66.5 | 632 ± 93.2 | **0.01** |
| HDL-Z diameter (nm) | 8.30 ± 0.06 | 8.29 ± 0.07 | 0.24 |
| HDL-P (µmol/L) | 25.2 ± 2.78 | 24.2 ± 2.97 | **0.008** |
| LargeHDL-P (µmol/L) | 0.26 ± 0.03 | 0.27 ± 0.03 | **0.02** |
| MediumHDL-P (µmol/L) | 9.52 ± 1.19 | 8.94 ± 0.98 | **<0.001** |
| SmallHDL-P (µmol/L) | 15.4 ± 2.37 | 15 ± 2.63 | 0.23 |
| s/t-VLDL-P ratio | 0.86 ± 0.02 | 0.87 ± 0.02 | 0.18 |
| s/t-LDL-P ratio | 0.58 ± 0.04 | 0.60 ± 0.04 | **<0.001** |
| s/t-HDL-P ratio | 0.61 ± 0.04 | 0.62 ± 0.05 | 0.14 |

*Abbreviations*: C, cholesterol; HDL, high-density lipoprotein; HDL-P, high-density lipoprotein particles; IDL, intermediate-density lipoprotein; LDL, low-density lipoprotein; LDL-P, low-density lipoprotein particles; P, particle concentration; SD, standard deviation; TG, triglycerides; VLDL, very-low-density lipoprotein; VLDL-P, very-low-density lipoprotein particles; Z, mean particle diameter; s/t-P, small-to-total particle-number ratios.

**Table S3. Comparison of glycoprotein variables between children with metabolically healthy obesity (MHO) and metabolically unhealthy obesity (MUO).** Data are presented as the mean ± standard deviation (SD). Wilcoxon rank-sum test was used to compare groups. The resulting *p*-values were corrected for multiple testing using the Benjamini–Hochberg false discovery rate (FDR) procedure. Bold values indicate statistical significance (FDR-adjusted q-value < 0.05).

| Glycoprotein variables | MHO (n=126) | MUO (n=141) | FDR q-values |
|---|---|---|---|
| Glyc-A (µmol/L) | 586 ± 64.7 | 675 ± 97 | **<0.001** |
| Glyc-B (µmol/L) | 313 ± 33.2 | 341 ± 34.6 | **<0.001** |
| Glyc-F (µmol/L) | 194 ± 22.9 | 222 ± 44 | **<0.001** |
| H/W Glyc-A | 14.3 ± 1.61 | 15.9 ± 1.83 | **<0.001** |
| H/W Glyc-B | 3.95 ± 0.42 | 4.29 ± 0.44 | **<0.001** |

*Abbreviations:* Glyc-A, glycoprotein A; Glyc-B, glycoprotein B; Glyc-F, glycoprotein F; H/W Glyc-A, height-to-width ratio of glycoprotein A; H/W Glyc-B, height-to-width ratio of glycoprotein B.

**Table S4. Comparison of low-molecular-weight metabolite (LMWM) profiles of children with metabolically healthy obesity (MHO) and metabolically unhealthy obesity (MUO).** Data are presented as the mean ± standard deviation (SD). Wilcoxon rank-sum test was used to compare groups. The resulting *p*-values were corrected for multiple testing using the Benjamini–Hochberg false discovery rate (FDR) procedure. Bold values indicate statistical significance (FDR-adjusted q-value < 0.05).

| LMWM variables | MHO (n=126) | MUO (n=141) | FDR q-values |
|---|---|---|---|
| Acetate (µmol/L) | 9.48 ± 5.82 | 7.84 ± 5.90 | **0.01** |
| Acetone (µmol/L) | 12.4 ± 8.97 | 10 ± 8.69 | **<0.001** |
| Alanine (µmol/L) | 264 ± 46.3 | 302 ± 52.7 | **<0.001** |
| Creatine (µmol/L) | 42.8 ± 21.4 | 45.8 ± 21.37 | 0.43 |
| Creatinine (µmol/L) | 24.3 ± 11.4 | 26.06 ± 10.6 | 0.25 |
| Glucose (µmol/L) | 3563 ± 424 | 3609 ± 479 | 0.39 |
| Glutamate (µmol/L) | 66.5 ± 15.9 | 73.4 ± 18.5 | **0.007** |
| Glutamine (µmol/L) | 355 ± 45.1 | 350 ± 41.3 | 0.29 |
| Glycine (µmol/L) | 191 ± 26.2 | 190 ± 25.4 | 0.87 |
| Histidine (µmol/L) | 70.7 ± 11.3 | 68.2 ± 12.5 | 0.12 |
| Isoleucine (µmol/L) | 27 ± 13.7 | 29 ± 14 | 0.32 |
| Lactate (µmol/L) | 294 ± 105 | 332 ± 110 | **0.009** |
| Leucine (µmol/L) | 75.9 ± 27 | 81 ± 27 | 0.09 |
| Proline (µmol/L) | 67.3 ± 27.3 | 75.8 ± 29.9 | **0.01** |
| Threonine (µmol/L) | 137 ± 55.2 | 129 ± 56.1 | 0.19 |
| Tyrosine (µmol/L) | 46.8 ± 10.3 | 49.2 ± 11 | **0.03** |
| Valine (µmol/L) | 145 ± 50 | 154 ± 51 | 0.06 |

**Table S5. Comparison of advanced lipidic profiles of children with metabolically healthy obesity (MHO) and metabolically unhealthy obesity (MUO).** Data are presented as the mean ± standard deviation (SD). Wilcoxon rank-sum test was used to compare groups. The resulting *p*-values were corrected for multiple testing using the Benjamini–Hochberg false discovery rate (FDR) procedure. Bold values indicate statistical significance (FDR-adjusted q-value < 0.05).

| **Lipidic variables** | **MHO (n=126)** | **MUO (n=141)** | **FDR q-values** |
|---|---|---|---|
| ARA+EPA (mmol/L) | 1.41 ± 0.19 | 1.47 ± 0.22 | **0.01** |
| DHA (mmol/L) | 0.14 ± 0.02 | 0.15 ± 0.02 | **0.005** |
| EC (mmol/L) | 2.52 ± 0.23 | 2.53 ± 0.28 | 0.97 |
| FC (mmol/L) | 1.49 ± 0.16 | 1.47 ± 0.18 | 0.41 |
| LA (mmol/L) | 3.52 ± 0.47 | 3.81 ± 0.76 | **0.002** |
| LPC (mmol/L) | 0.56 ± 0.05 | 0.57 ± 0.06 | 0.66 |
| PC (mmol/L) | 2.27 ± 0.21 | 2.27 ± 0.24 | 0.88 |
| PL (mmol/L) | 2.79 ± 0.29 | 2.72 ± 0.31 | 0.14 |
| PUFAs (mmol/L) | 11.7 ± 1.41 | 12 ± 1.54 | 0.11 |
| SFAs (mmol/L) | 7.87 ± 0.52 | 8 ± 0.58 | 0.10 |
| SM (mmol/L) | 0.83 ± 0.07 | 0.82 ± 0.07 | 0.14 |
| ω-3 (mmol/L) | 0.30 ± 0.05 | 0.31 ± 0.06 | 0.06 |
| ω-6+ω-7 (mmol/L) | 4.71 ± 0.61 | 4.91 ± 0.83 | **0.03** |
| ω-9 (mmol/L) | 2.95 ± 0.45 | 3.65 ± 0.92 | **<0.001** |

Abbreviations: ARA+EPA, Arachidonic acid + Eicosapentaenoic acid; DHA, Docosahexaenoic acid; EC, Esterified cholesterol; FC, Free cholesterol; LA, Linoleic acid; LPC, Lysophosphatidylcholine; PC, Phosphatidylcholine ; PL, Glycerophospholipids (except LPC); PUFA, Polyunsaturated fatty acids; SFA, Saturated fatty acids; SM, Sphingomyelin; ω3, omega-3 fatty acids; ω6+ω7, omega-6 and 7 fatty acids; ω9, omega-9 fatty acids.

**Table S6. Comparison of anthropometric, clinical, biochemical, and NMR-metabolomics variables between the two clusters derived from unsupervised k-means clustering analysis.** Continuous variables are presented as the mean ± standard deviation (SD), except for systolic and diastolic blood pressure percentiles, which are presented as median (interquartile range, IQR). Sex is reported as n (%). We used the Wilcoxon rank-sum test to compare continuous variables between the two clusters, and the chi-square test to compare categorical variables. P-values were corrected for multiple testing using the Benjamini-Hochberg FDR method. Bolded values indicate statistical significance (FDR-adjusted q-value < 0.05).

| | **Cluster 1 (n=150)** | **Cluster 2 (n=117)** | **FDR q-values** |
|---|---|---|---|
| **Anthropometric and clinical characteristics** | | | |
| BMI (kg/m$^2$) | 25.8 ± 2.7 | 25.8 ± 2.8 | 0.65 |
| BMI z-score | 2.58 ± 0.45 | 2.57 ± 0.44 | 0.73 |
| Age (years) | 10.8 ± 1.6 | 10.8 ± 1.6 | 0.88 |
| Sex (n; %) | | | 0.93 |
| Male | 81 (54%) | 64 (55%) | |
| Female | 69 (46%) | 53 (45%) | |
| Weight (kg) | 57.8 ± 12.5 | 57.2 ± 12.7 | 0.70 |
| Height (m) | 1.49 ± 0.11 | 1.48 ± 0.10 | 0.87 |
| Waist/Height | 0.57 ± 0.04 | 0.58 ± 0.04 | 0.10 |
| Fat Mass Index | 10.4 ± 2.2 | 10.5 ± 2.0 | 0.81 |
| Fat Free Mass Index | 15.4 ± 1.88 | 15.3 ± 1.47 | 0.47 |
| SBP (percentile) | 68 (45–86) | 69 (47–85) | 0.74 |
| DBP (percentile) | 45 (31–59) | 47 (34–66) | 0.31 |
| **Biochemical Parameters for conventional lipid profile** | | | |
| TC (mg/dL) | 148 ± 20 | 181 ± 26 | **<0.001** |
| TG (mg/dL) | 62.1 ± 18.7 | 111 ± 42 | **<0.001** |
| HDL-C (mg/dL) | 53 ± 11 | 51.2 ± 11.5 | 0.29 |

| | | | |
|---|---|---|---|
| LDL-C (mg/dL) | 82.3 ± 17.7 | 107 ± 25 | **<0.001** |
| Fasting glucose (mg/dL) | 83.7 ± 5.33 | 84.2 ± 6.20 | 0.73 |
| Fasting insulin (µIU /mL) | 12.6 ± 7.24 | 15.6 ± 8.93 | **0.002** |
| HOMA-IR | 2.64 ± 1.54 | 3.29 ± 2 | **0.002** |
| MetScore | 3.57 ± 3.04 | 5.96 ± 3.67 | **<0.001** |
| **NMR-derived variables** | | | |
| VLDL-C (mg/dL) | 10 ± 3 | 17.3 ± 6.1 | **<0.001** |
| VLDL-TG (mg/dL) | 44.1 ± 11.1 | 69.1 ± 24.7 | **<0.001** |
| TG/C-VLDL | 4.53 ± 0.73 | 4.03 ± 0.49 | **<0.001** |
| IDL-C (mg/dL) | 5.42 ± 1.24 | 8.09 ± 1.88 | **<0.001** |
| IDL-TG (mg/dL) | 6.87 ± 1.05 | 9.14 ± 1.41 | **<0.001** |
| TG/C-IDL | 1.30 ± 0.17 | 1.16 ± 0.14 | **<0.001** |
| LDL-C (mg/dL) | 100 ± 12 | 115 ± 17 | **<0.001** |
| LDL-TG (mg/dL) | 8.32 ± 1.58 | 11.6 ± 2.32 | **<0.001** |
| TG/C-LDL | 0.08 ± 0.01 | 0.10 ± 0.02 | **<0.001** |
| HDL-C (mg/dL) | 49.3 ± 6.2 | 49.4 ± 6.3 | 0.93 |
| HDL-TG (mg/dL) | 11 ± 1 | 14 ± 2 | **<0.001** |
| TG/C-HDL | 0.23 ± 0.04 | 0.29 ± 0.07 | **<0.001** |
| VLDL-Z diameter (nm) | 42.2 ± 0.2 | 42.2 ± 0.2 | 0.75 |
| VLDL-P (nmol/L) | 31.9 ± 7.7 | 50.5 ± 17.6 | **<0.001** |
| LargeVLDL-P (nmol/L) | 1.02 ± 0.28 | 1.42 ± 0.42 | **<0.001** |
| MediumVLDL-P (nmol/L) | 3.2 ± 1.1 | 5.3 ± 2.1 | **<0.001** |
| SmallVLDL-P (nmol/L) | 27.7 ± 6.6 | 43.8 ± 15.6 | **<0.001** |
| LDL-Z diameter (nm) | 21 ± 0.2 | 21 ± 0.2 | 0.96 |
| LDL-P (nmol/L) | 979 ± 116 | 1151 ± 159 | **<0.001** |
| LargeLDL-P (nmol/L) | 168 ± 18 | 183 ± 25 | **<0.001** |
| MediumLDL-P (nmol/L) | 227 ± 55 | 303 ± 75 | **<0.001** |
| SmallLDL-P (nmol/L) | 583 ± 61 | 664 ± 84 | **<0.001** |
| HDL-Z diameter (nm) | 8.31 ± 0.06 | 8.27 ± 0.06 | **<0.001** |
| HDL-P (µmol/L) | 24 ± 3 | 25 ± 3 | **<0.001** |
| LargeHDL-P (µmol/L) | 0.26 ± 0.03 | 0.26 ± 0.03 | 0.45 |
| MediumHDL-P (µmol/L) | 9.21 ± 1.15 | 9.22 ± 1.09 | 0.94 |
| SmallHDL-P (µmol/L) | 14.5 ± 2.4 | 16.2 ± 2.3 | **<0.001** |
| s/t-VLDL-P ratio | 0.87 ± 0.02 | 0.86 ± 0.02 | 0.22 |
| s/t-LDL-P ratio | 0.60 ± 0.04 | 0.58 ± 0.04 | **<0.001** |
| s/t-HDL-P ratio | 0.60 ± 0.05 | 0.63 ± 0.04 | **<0.001** |
| Glyc-A (µmol/L) | 581 ± 62 | 700 ± 85 | **<0.001** |
| Glyc-B (µmol/L) | 317 ± 34 | 341 ± 36 | **<0.001** |
| Glyc-F (µmol/L) | 190 ± 21 | 234 ± 41 | **<0.001** |
| H/W Glyc-A | 14.3 ± 1.61 | 16.2 ± 1.70 | **<0.001** |
| H/W Glyc-B | 4 ± 0.43 | 4.29 ± 0.46 | **<0.001** |
| Acetate (µmol/L) | 8.93 ± 6.26 | 8.20 ± 5.42 | 0.53 |
| Acetone (µmol/L) | 11.9 ± 9.1 | 10.3 ± 8.6 | 0.06 |
| Alanine (µmol/L) | 273 ± 50 | 298 ± 54 | **<0.001** |
| Creatine (µmol/L) | 41.6 ± 21.5 | 48.0 ± 20.8 | 0.06 |
| Creatinine (µmol/L) | 23.8 ± 11.1 | 27.2 ± 10.6 | **0.02** |
| Glucose (µmol/L) | 3617 ± 383 | 3550 ± 530 | 0.47 |
| Glutamate (µmol/L) | 68.2 ± 15.7 | 72.7 ± 19.7 | 0.07 |
| Glutamine (µmol/L) | 358 ± 44 | 345 ± 41.2 | **0.02** |
| Glycine (µmol/L) | 192 ± 26 | 189 ± 25 | 0.47 |
| Histidine (µmol/L) | 69.6 ± 11.7 | 69.2 ± 12.5 | 0.86 |
| Isoleucine (µmol/L) | 26 ± 13 | 31 ± 14 | **0.01** |
| Lactate (µmol/L) | 295 ± 102 | 339 ± 113 | **0.01** |

| | | | |
|---|---|---|---|
| Leucine (μmol/L) | 72.5 ± 27.4 | 86.3 ± 24.7 | **<0.001** |
| Proline (μmol/L) | 68.9 ± 29.6 | 75.5 ± 27.8 | 0.12 |
| Threonine (μmol/L) | 135 ± 58.8 | 130 ± 51.8 | 0.22 |
| Tyrosine (μmol/L) | 46.8 ± 11.1 | 49.7 ± 10.1 | **0.01** |
| Valine (μmol/L) | 141 ± 51.2 | 161 ± 45 | **<0.001** |
| ARA+EPA (mmol/L) | 1.35 ± 0.19 | 1.56 ± 0.16 | **<0.001** |
| DHA (mmol/L) | 0.14 ± 0.02 | 0.16 ± 0.02 | **<0.001** |
| EC (mmol/L) | 2.40 ± 0.21 | 2.69 ± 0.22 | **<0.001** |
| FC (mmol/L) | 1.39 ± 0.13 | 1.58 ± 0.15 | **<0.001** |
| LA (mmol/L) | 3.27 ± 0.40 | 4.20 ± 0.53 | **<0.001** |
| LPC (mmol/L) | 0.54 ± 0.04 | 0.60 ± 0.04 | **<0.001** |
| PC (mmol/L) | 2.17 ± 0.19 | 2.40 ± 0.20 | **<0.001** |
| PL (mmol/L) | 2.63 ± 0.28 | 2.91 ± 0.25 | **<0.001** |
| PUFAs (mmol/L) | 11.1 ± 1.1 | 12.9 ± 1.2 | **<0.001** |
| SFAs (mmol/L) | 7.78 ± 0.54 | 8.13 ± 0.53 | **<0.001** |
| SM (mmol/L) | 0.80 ± 0.06 | 0.87 ± 0.07 | **<0.001** |
| ω-3 (mmol/L) | 0.28 ± 0.04 | 0.34 ± 0.05 | **<0.001** |
| ω-6+ω-7 (mmol/L) | 4.34 ± 0.50 | 5.42 ± 0.55 | **<0.001** |
| ω-9 (mmol/L) | 2.93 ± 0.49 | 3.81 ± 0.88 | **<0.001** |

*Abbreviations*: BMI, body mass index; SBP, systolic blood pressure; DBP, diastolic blood pressure; TC, total cholesterol; TG, triglycerides; HDL-C, high-density lipoprotein cholesterol; LDL-C, low-density lipoprotein cholesterol; VLDL-C, very low-density lipoprotein cholesterol; HOMA-IR, Homeostasis Model Assessment of Insulin Resistance; s/t-P, small-to-total particle-number ratios; Glyc-A, glycoprotein A; Glyc-B, glycoprotein B; Glyc-F, glycoprotein F; H/W Glyc-A, height-to-width ratio of glycoprotein A; H/W Glyc-B, height-to-width ratio of glycoprotein B; ARA+EPA, Arachidonic acid + Eicosapentaenoic acid; DHA, Docosahexaenoic acid; EC, Esterified cholesterol; FC, Free cholesterol; LA, Linoleic acid; LPC, Lysophosphatidylcholine; PC, Phosphatidylcholine ; PL, Glycerophospholipids (except LPC); PUFA, Polyunsaturated fatty acids; SFA, Saturated fatty acids; SM, Sphingomyelin; ω3, omega-3 fatty acids; ω6+ω7, omega-6 and 7 fatty acids; ω9, omega-9 fatty acids.

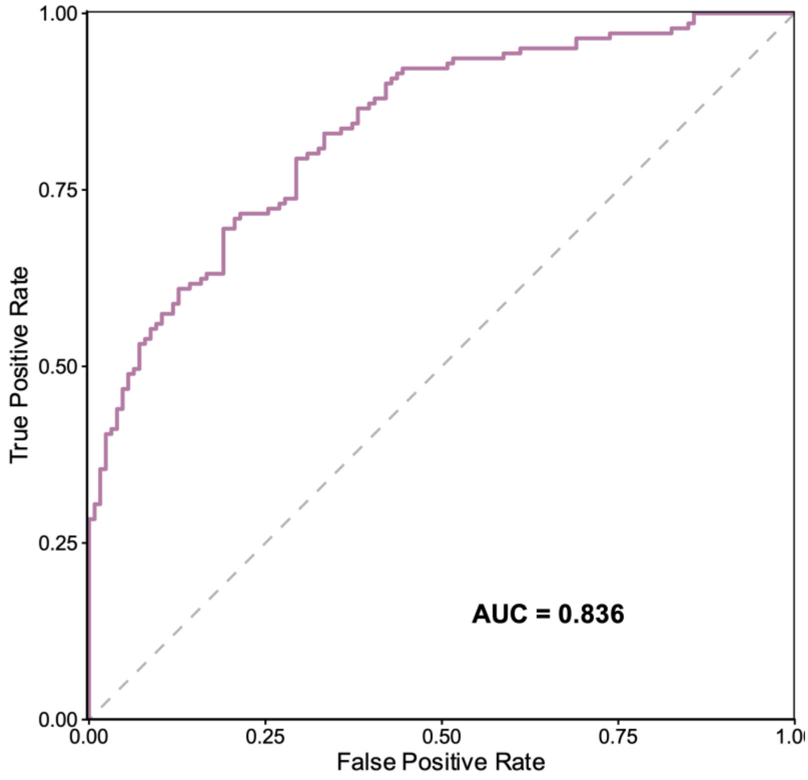

**Figure S1. Receiver operating characteristic (ROC) curve for the PLS-DA model discriminating MUO from MHO.** ROC curve was generated from cross-validated out-of-fold predictions across 10-fold cross-validation. The area under the curve (AUC) was 0.83. The diagonal dashed line represents the line of no discrimination (AUC = 0.5).
